# Perceived changes in competence, relatedness, and autonomy reported by mothers since joining a mom-centered digital community

**DOI:** 10.1101/2024.07.09.24310140

**Authors:** Katherine E. McManus-Shipp, Christiana M. Field, Sandesh Bhusal, Cindy-Lee Dennis, Molly E. Waring

**Author notes:** <u>Corresponding author:</u> Molly E. Waring, PhD, Department of Allied Health Sciences, University of Connecticut, 358 Mansfield Rd, Unit 1101, Storrs, CT 06269.

## Abstract

**Background:** Motherhood can profoundly challenge individuals’ well-being. Social media and other digital platforms are promising modalities for reaching and supporting mothers with evidence-based psychoeducation and connection to peers. However, much is unknown about how mothers perceive these online peer communities and their impact on health and well-being.

**Purpose:** To describe mothers’ perceptions of the impact of exposure to and engagement with a mom-centered digital community (Momwell) on their well-being.

**Methods:** Pregnant persons and mothers exposed to Momwell psychoeducational content and community related to motherhood via social media, podcast, or blog completed an online survey (N=569). Participants reported several perceived changes related to competence, relatedness/connection, and autonomy in decision-making since joining the Momwell community by rating their agreement with a series of questions.

**Results:** All but two participants identified as mothers; 45% were either pregnant or within 12 months postpartum. The majority agreed with statements about perceived changes in their lives, well-being, and feelings since joining the Momwell community (82-97%). All participants reported positive changes related to their sense of competence, 99% reported positive changes related to relatedness, and 97% reported positive changes related to autonomy.

**Conclusion:** Exposure to psychoeducational content related to motherhood and maternal mental health and peer engagement within a mom-centered community can enhance maternal well-being through positive changes in competence, relatedness, and autonomy.

## Introduction

The birth of a child dramatically changes one’s life and routines, and many mothers feel pressured to live up to societal expectations of motherhood or struggle to balance life roles and responsibilities, which can negatively impact their mental health [1,2] and well-being [3,4].

Believing in their ability to parent, and experiencing a sense of belonging, and feeling empowered to make decisions can improve overall well-being. This relates to one of the most common and well-researched theories on well-being known as Self-Determination Theory (SDT) [5]. This theory states that individuals have three basic needs that influence their well-being: (1) *competence*, or confidence and belief in one’s abilities, (2) *relatedness*, or the need to feel connected to and supported by others and experience a sense of belonging, and (3) *autonomy*, or the feeling that one has a choice in their decision-making, behavior, and attitudes. SDT is frequently integrated into the parenting literature, where it is often applied to infant feeding practices [6,7] and improving child socioemotional and behavioral outcomes [8]. However, fewer studies have applied this theory to maternal well-being during the early stages of parenting [9]. Given the evidence to support the role of competence, relatedness, and autonomy in fostering healthy psychosocial functioning and well-being in children, understanding the role these concepts play in facilitating the well-being of mothers as they navigate the challenges of parenting is needed.

Mothers seek information and support from peers, often on social media or other digital platforms [3,4,10,11]. As the majority of women of childbearing age in the United States [12–14] and Canada [15] use social media, and an increasing proportion listen to podcasts [16], these digital media may be a way to connect pregnant persons and mothers with psychoeducation and support from peers. However, little is known about how pregnant persons and mothers perceive the information and support they receive from online peer communities and how it impacts their lives. Specifically, it is unclear how mothers utilize information regarding mental health, how the peer community influences their perceptions and confidence as mothers, and how such information impacts their willingness to identify and prioritize their own mental health and physical needs.

To address this gap, we conducted a research study to survey perinatal persons and mothers who followed and engaged with a mom-centered digital community, Momwell [17]. Momwell offers a mom-centered model of care that seeks to educate, empower, and support mothers (momwell.com). Founded and led by a psychotherapist based in Canada, Momwell provides evidence-based psychoeducational content about motherhood, parenting, and maternal mental health through its social media feeds (Instagram, Facebook, and TikTok), weekly podcasts, and blogs with posts corresponding to the podcast episodes. Based on SDT, exposure to and engagement with Momwell content and the peer community may build competence by providing psychoeducation and resources that mothers can apply daily. Online peer communities like Momwell can enhance relatedness by creating opportunities for connection with other mothers and encouraging partner and family involvement in childcare. Further, exposure to and engagement with Momwell psychoeducational content and peer community may strengthen autonomy by supporting mothers in making evidence-informed decisions related to their physical and mental health and parenting practices.

In this report, we describe maternal perceived changes in knowledge, attitudes, and competence related to motherhood and mental health since joining the Momwell community. We also describe perceived changes related to the SDT domains of competence, relatedness, and autonomy.

## Methods

In the summer and early fall of 2023, we recruited perinatal persons and mothers who were exposed to or engaged with Momwell via social media or other digital platforms to complete an online survey. A detailed description of the study design is available elsewhere [17]. Briefly, we collaborated with the Momwell team to post recruitment messages on social media, in their email newsletter, and via a podcast advertisement. Interested individuals clicked a link to complete an eligibility screener via Qualtrics. Eligible individuals were 18 years or older, a member of the Momwell community (i.e., followed Momwell on Instagram, Facebook, or TikTok, listened to their podcast, read their blog, or received their email newsletters), comfortable participating in the study in English, and able and willing to provide informed consent. Individuals were recruited to participate in two cohorts where Cohort 1 completed a longer survey and Cohort 2 completed a shorter survey. Additional eligibility criteria for Cohort 1 were (1) perinatal (e.g., currently pregnant and/or within 12 months postpartum) and/or identified as the mother of at least one child under the age of 18 who lived with them at least part-time, and (2) living in Canada or the United States. We employed several strategies to ensure the eligibility and validity of enrolled participants (for details, see [17]). Participants in Cohort 1 who completed the longer survey received a gift card ($15 USD or $20 CAD). Cohort 2 participants who completed the shorter survey were entered into a lottery in which one in every 30 participants was randomly selected to receive a gift card ($60 USD or $80 CAD). The University of Connecticut Institutional Review Board (IRB) approved this study.

Participants reporting exposure to psychoeducation content on the Momwell platform were asked how recently they started following Momwell on each of their digital platforms (within the past 7 days, at least 7 days ago but less than 3 months ago, at least 3 months ago but less than 6 months ago, at least 6 months ago but less than 12 months ago, and at least 12 months ago). Participants who started following Momwell within the past 7 days were considered new community members and were not asked questions about changes in their lives and well-being.

Participants reported the extent to which they agreed with several statements about changes to their lives since joining the Momwell community (e.g., “I feel more aware of the signs and symptoms of mental health conditions”, “I feel more confident in my approach to parenting”). Response options were “strongly disagree”, “disagree”, “agree”, and “strongly agree”. We calculated the proportion of participants who responded “agree” or “strongly agree” for each item. We mapped these questions onto the three domains of SDT (Table 1). We calculated changes related to each domain as the highest level of agreement with questions within that domain. For the reversed item, we reversed coding and considered “disagree” to be “agree” when calculating the composite variable for competence. Internal consistency of these composite variables was good (Cronbach’s α = 0.77, 0.75, and 0.70, respectively, for the competence, relatedness, and autonomy scores).

**Table 1:**
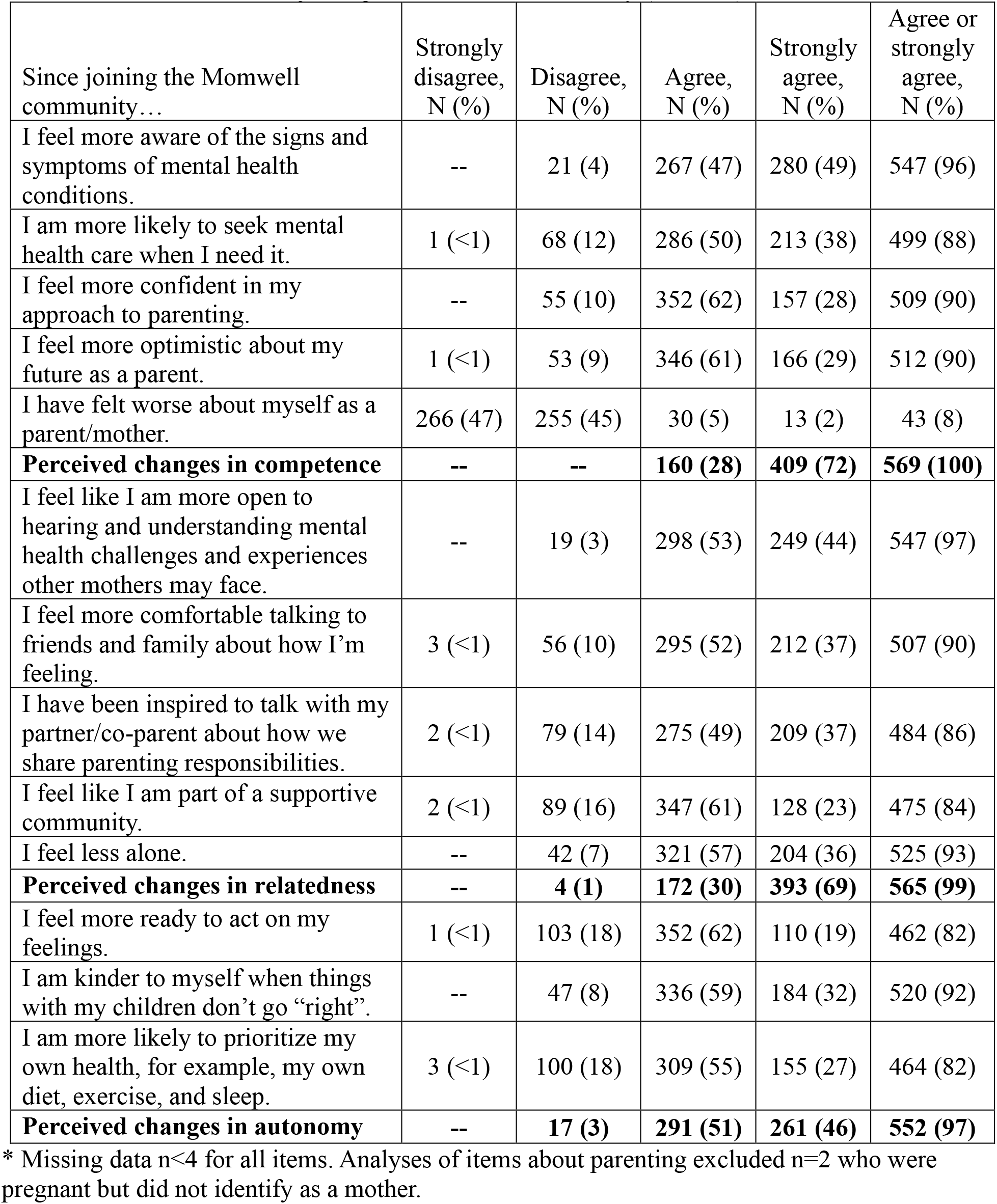
Perceived changes in knowledge, beliefs, and competence related to motherhood and maternal mental health since joining the Momwell community (N=569*)

Participants reported whether they were pregnant or postpartum (i.e., within 12 months of giving birth) and whether they identified as a mother of one or more children aged 0-17 years who lived with them at least part-time. All but two participants identified as mothers (they were pregnant) and thus in this study we refer to participants as “mothers”. Participants also reported their gender, country of residence, educational attainment, employment status, and whether they self-identified as someone who was a visible minority [18]. Participants reported how difficult it has been for them in the past 30 days to pay for usual household expenses (e.g., food, rent, car payments, medical expenses) as a measure of financial strain [19]. Participants in Cohort 1 were further asked whether they had a parenting partner. Characteristics of the full sample are described elsewhere [17].

## Statistical analyses

The eligibility screener and surveys were administered in Qualtrics (Qualtrics LLC, Provo, UT). We used Research Electronic Data Capture (REDCap) for participant tracking [20]. Analyses were conducted in SAS 9.4 (SAS Institute, Inc., Cary, NC). After excluding n=15 who joined the Momwell community in the past 7 days who were not asked the questions of interest, our analytic sample included 569 mothers. We described the characteristics of the analytic sample and then participant responses to questions about changes they have made since they joined the Momwell community. We also described the proportion of participants who perceived changes related to the three domains of SDT: competence, relatedness, and autonomy.

We conducted multiple sensitivity analyses. As one item asked about communication with their partner/co-parent, we repeated the analysis limited to (1) participants who reported that their children lived with 2 parents in the same household (n=542), and (2) mothers in Cohort 1 were reported that they had a parenting partner (n=280; only participants in Cohort 1 were asked whether they had a parenting partner). Twelve participants selected the same response for all 13 items (n=8 all “agree”; n=4 all “strongly agree”). As one item was reverse worded, such that agreeing or strongly agreeing represented a negative change since joining the Momwell community, this response pattern may represent inattention to the survey items (i.e., straight-lining). We conducted a sensitivity analysis in which we excluded these participants (N=557).

Finally, as our interest was in the perceived impact of the free psychoeducational content and peer community, we conducted a sensitivity analysis limited to participants who had never purchased for-fee services (i.e., guided journal or other self-paced tool, course or workshop, therapy with Momwell therapist; n=423).

## Results

Participants in the analytic sample (N=569) were on average 34.2 (SD: 3.9) years old, 45% were perinatal (10% pregnant, 36% post-partum), 13% self-identified as a visible minority, 87% had a 4-year college/university degree or higher education, 49% were working full-time, and 43% reported some difficulty in paying for basic expenses. Most (76%) participants had joined the Momwell community more than 12 months ago, 16% had joined at least 6 months but less than 12 months ago, and 8% had joined in the past 6 months.

Most participants agreed or strongly agreed with statements about perceived changes in their lives, well-being, and feelings since joining the Momwell community (Table 1). Overall, 82% reported that they are now more likely to prioritize their own health and 96% indicated they felt more aware of the signs and symptoms of mental health problems (Table 1). In terms of changes related to the SDT domains, 100% reported an improved sense of competence, 99% reported increased feelings of relatedness to other mothers, and 97% indicated enhanced autonomy (Figure 1). Results from sensitivity analyses were very similar to the main results (Supplemental Table 1).

## Discussion

We found the majority of mothers reported increased competence, relatedness, and autonomy since joining the Momwell community. Participants expressed that they had made several positive changes related to their physical and mental health, self-efficacy as mothers, and relationships with partners/co-parents and children since joining the Momwell community. Importantly, 96% reported feeling more aware of the signs and symptoms of mental health problems. Such results are promising given that poor mental health literacy is a well-established barrier to help-seeking [2] and effective mental health literacy interventions are said to improve both knowledge and help-seeking [21]. Additionally, 88% reported being more likely to seek mental health care when they needed it. Given that many women experience elevated post-partum mood symptoms yet do not seek care [22], our results are particularly encouraging and suggest that utilizing mom-centered digital communities may be an important promotor of help-seeking behavior. Further, participants reported they felt more confident and optimistic in their parenting approach since joining the Momwell community, suggesting that exposure to and engagement with digital platforms may increase maternal competence and confidence. These results align with a recent systematic review that found that participation in digital parenting interventions increased parenting self-efficacy [23].

Participants reported an increased sense of relatedness where they felt they were part of a community and were more willing to communicate with their partners about their needs. This finding suggests that following a mom-centered account on social media or other digital platforms can foster a sense of community and belonging and may increase maternal perceived support. Previous studies have demonstrated the benefits of peer support on perinatal and maternal mental health [24,25], and the current findings hold important implications for partner support and potentially improving maternal relationship satisfaction and co-parenting outcomes. This is critical given perceived partner support is repeatedly associated with positive maternal and infant outcomes postpartum [26] and maternal mental health outcomes [27].

Most participants reported increased autonomy, particularly feeling ready to act on their feelings, treat themselves with self-compassion, and prioritize their health. While mothers of young children often do not prioritize their own health, more than 80% of mothers in our study reported that they were more likely to prioritize their health since joining the Momwell community. Given the importance of diet, physical activity, and sleep to long-term cardiometabolic health [28], our finding that mothers feel more empowered to engage in health behaviors after engaging with a digital mom-centered community is clinically relevant.

Consistent with our results, a recent study of mothers with children under 3 years found that the two most commonly reported external facilitators to engaging in self-care were partners and encouragement from others [29]. Further, we found that 92% of mothers reported being kinder to themselves during challenging times with their children since joining the Momwell community. As self-compassion has been associated with more positive views on prioritizing one’s own health behaviors among mothers [30], online communities such as Momwell may also increase mothers’ prioritization of their health through increased self-compassion. Future studies should investigate ways in which exposure to or engagement with social media content and peer communities impacts maternal health behaviors and perceived autonomy to make health-promoting decisions.

This study has limitations. Common to much survey research, our sample consisted of members of the target population who volunteered to participate in our study. Thus, perceived changes since joining the Momwell community may overrepresent the experiences of those for whom Momwell has positively influenced their lives. Additionally, our sample likely overrepresents Momwell followers who are more engaged on social media and thus more likely to see our recruitment messages [31,32]. We reported perceived changes in maternal thoughts, feelings, and actions since joining the Momwell community measured by a single item per domain. While these questions have high face validity, social desirability bias in responding is possible. Future research studies could use longer, validated psychological scales measuring each domain. It is likely that participants experienced many life changes since they joined the Momwell community, and these experiences and other factors likely also influenced changes in competence, relatedness, and autonomy related to maternal well-being over this time period.

Future research could extend these results by incorporating qualitative perspectives from mothers to provide additional context to our findings, for example, asking mothers to describe what content or interaction in the community had the biggest positive impact on them. This line of inquiry would allow researchers to more precisely pinpoint influential psychoeducational content to improve mothers’ sense of competence, relatedness, and autonomy. Social media content platforms and providers can then incorporate these research insights. Further, future research should explore how competence, relatedness, and autonomy change over time, as mothers join mom-centered digital communities, consume psychoeducational content, and engage with peers. It would also be worthwhile to investigate the relationships between exposure and engagement, SDT variables, and mental health outcomes as a marker of well-being and to identify for whom engagement in these platforms is most beneficial. Findings from such studies could inform the development of clinical practice guidelines that can assist healthcare providers in recommending online communities and information sources to improve health outcomes. Finally, policymakers should recognize the value of online support initiatives that expand access to digital health resources and aid healthcare providers in integrating online psychoeducation resources into their standard clinical care.

This study contributes to our understanding of the benefits mothers perceive from health information and peer support they receive through social media and other digital platforms such as podcasts and blogs. Our findings suggest that exposure to and engagement with digital mom-centered psychoeducational content and peer community focused on perinatal and maternal mental health and parenting may enhance maternal competence, relatedness, and autonomy, thus influencing overall well-being. SDT provides a useful framework for understanding the impact on mothers of exposure to and engagement with psychoeducational content and community provided through social media and other digital platforms.

## Data Availability

Most of the data presented in this paper are available online at OpenICPSR (https://doi.org/10.3886/E204321V3). All data presented in this paper are available upon reasonable request to the first author.

## Acknowledgements

This study was funded by a grant from the Daymark Foundation (PI: Waring). Vani Jain of the Daymark Foundation and Erica Djossa of Momwell provided input into the design of this study, particularly by providing input into the research questions and concepts to be measured, and Erica Djossa and Momwell staff helped to develop recruitment messages (which were then approved by the University of Connecticut IRB) and disseminated these messages to their followers/subscribers. The Daymark Foundation and Momwell had no additional role in data collection or results reporting.

## Tables and Figures

### Online Supplemental Appendix

**Supplemental Table 1:**
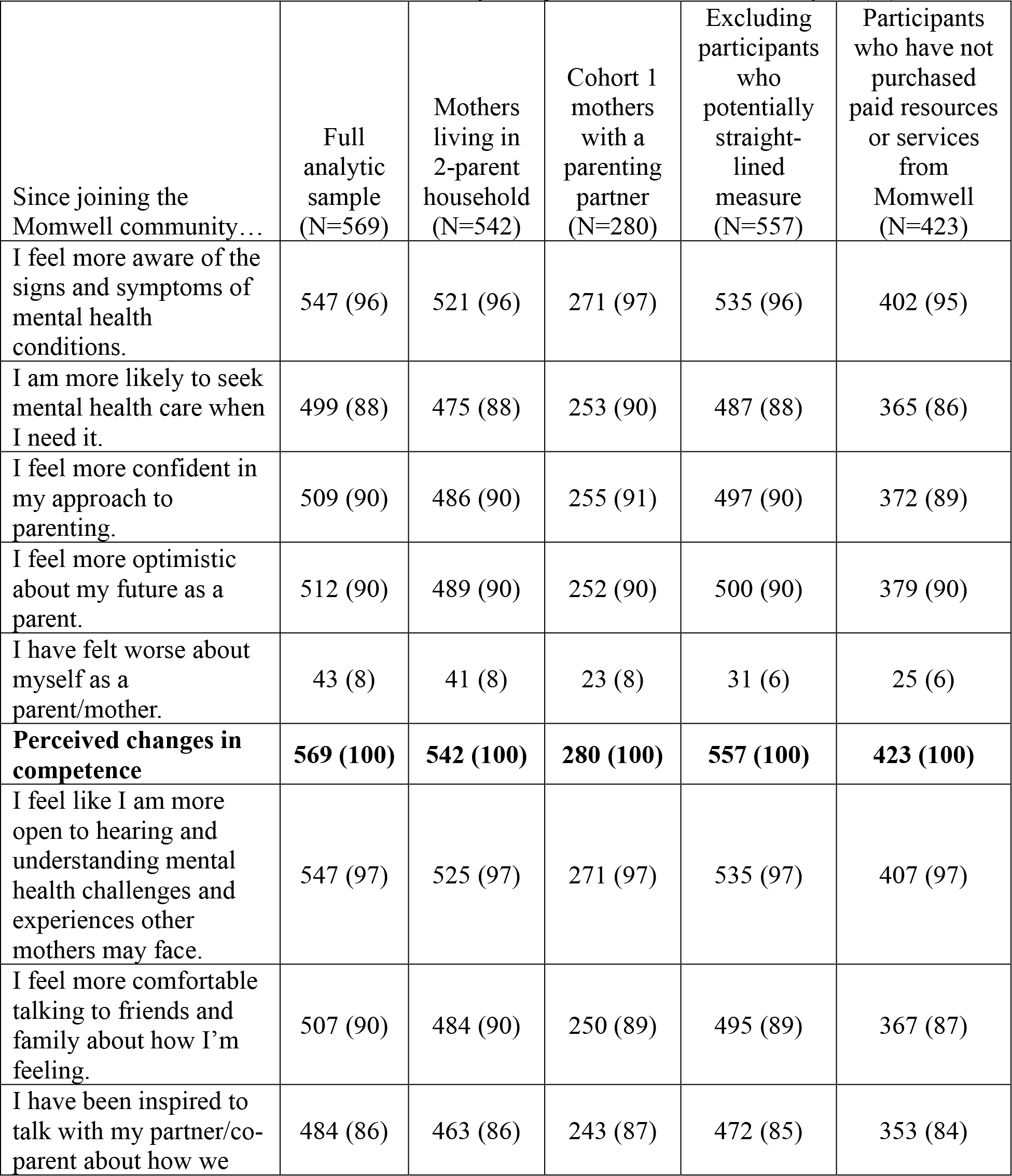
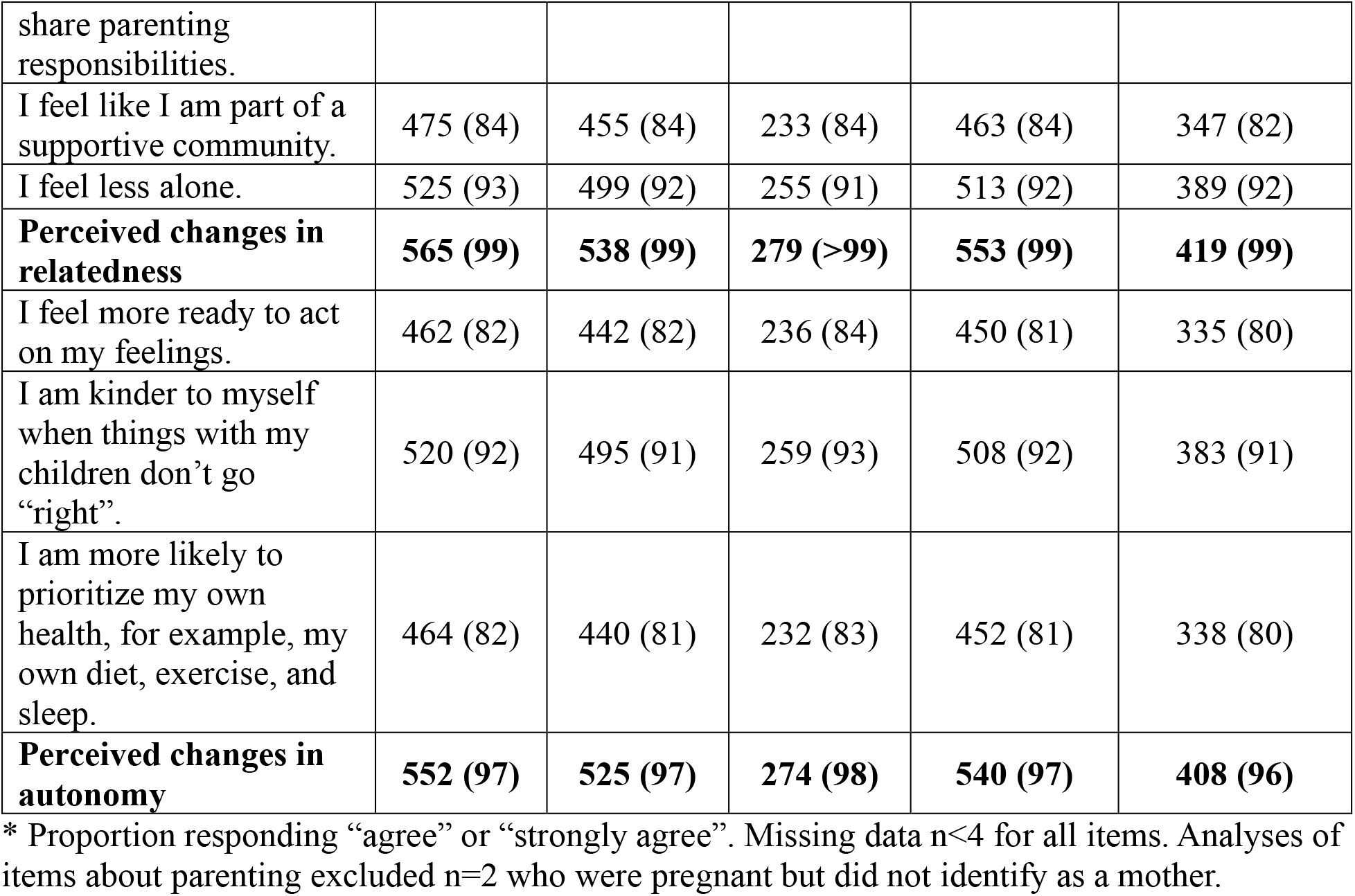
Perceived changes in knowledge, beliefs, and competence related to motherhood and maternal mental health since joining the Momwell community, N (%)*

